# NEAT (NicotinE As Treatment) Trial: Protocol of a randomised controlled trial of vaporised nicotine products compared with nicotine replacement therapy following discharge from residential withdrawal services

**DOI:** 10.1101/2022.05.15.22275118

**Authors:** B Bonevski, JL Rich, E Skelton, J Garfield, AL Baker, C Segan, C Gartner, N Walker, R Borland, M Daglish, A Dunlop, C Oldmeadow, L Bauld, C Bullen, N Ezard, R McCrohan, D Jacka, S White, DI Lubman, V Manning

**Author notes:** **Corresponding author:** Professor Billie Bonevski, Flinders Health and Medical Research Institute, College of Medicine and Public Health, Flinders University, Bedford Park, South Australia, 5048, Australia.

## Abstract

**Background:** Tobacco smoking rates in alcohol and other drug (AOD) treatment settings is much higher than in the general Australian population. As a result, people seeking treatment for AOD use experience a greater tobacco-related burden of illness. Attempts to reduce smoking rates in AOD treatment consumers have failed to identify smoking cessation strategies with long term effectiveness. The primary aim of this study is to examine the effectiveness of nicotine vaporised products (NVPs) or nicotine replacement therapy (NRT)on self-reported 6 months continuous abstinence at the 9-month follow-up (6 months following end-of 12 weeks of nicotine treatment) for people leaving smoke-free residential withdrawal treatment. Both groups will also receive Quitline telephone counselling. Secondary outcomes and process measures will also be collected.

**Methods:** A two-arm, single-blinded, parallel-group randomised trial with a 6-month post-intervention follow-up (9 months following baseline) will be conducted. The setting is five residential and inpatient government-funded AOD withdrawal units across three cities in three states of Australia (New South Wales, Queensland, Victoria). Participants will be service users aged 18 years or over who smoked at least 10 cigarettes per day, interested in quitting in the next 30 days and have capacity to give informed consent. Research assistants will recruit participants during intake, who then complete a baseline survey, will be randomised to a condition, and receive their first Quitline call during AOD treatment. At discharge, all participants receive a discharge pack containing either NVPs or NRT, depending on condition allocation.

**Discussion:** This is the first study we know of that will be testing intervening with a tobacco smoking cessation approach during the transition phase from AOD treatment to community. From a public health perspective, this approach has the potential to have tremendous reach into a priority population for smoking cessation.

**Trial registration:** Australian and New Zealand Clinical Trials Registry (ACTRN12619001787178)

## INTRODUCTION

Up to 95% of people in alcohol and other drug (AOD) treatment in Australia smoke tobacco. [1, 2] This is in comparison with population-wide smoking rates in Australia of 12%. [3] People with AOD dependence report higher rates of moderate and heavy nicotine dependence and smoke more cigarettes per day than the general population. [4-6] As a result, people seeking treatment for AOD use experience a greater tobacco-related burden of illness. [7, 8] Tobacco-related deaths are higher among people who have been treated for AOD dependence than the general population. [9]

People in AOD treatment are greatly interested in quitting smoking. [5, 10] When given evidence-based smoking cessation interventions during AOD treatment, studies have shown that short-term smoking cessation is possible and safe. [11, 12] Recent Australian research suggests that smoking cessation can improve treatment success for other drug use. [13] However, an ongoing challenge is sustaining abstinence from smoking in the longer term (at least 6 months). [14, 15] High relapse rates in this population may be due to factors related to AOD dependence (e.g. smoking-related cues and triggers), lack of cessation support, and the high levels of smoking in their social network. [2, 16] Three large trials providing AOD treatment service consumers with behavioural support and nicotine replacement therapy (NRT) reported short-term abstinence (8-12 weeks) rates between 9-33% while in treatment, but most relapsed to smoking following discharge. [17-19] The heavy nicotine dependence amongst people with AOD dependence is an important contributor to this high relapse rate. [16] Although NRT can reduce withdrawal symptoms and cravings and aid cessation, research with people with AOD dependence indicates NRT is underutilised. [20]

Nicotine vaping products (NVPs) are a broad range of battery powered devices that deliver an aerosol of propylene glycol and/or glycerol, nicotine and flavours. [21] In Australia, access to liquid nicotine is available using the Personal Importation Scheme with a presecription from a General Practitioner for use as a smoking cessation tool. [22] Unlike combustible tobacco cigarettes, NVPs deliver nicotine in an inhalable form without burning tobacco. Like pharmaceutical NRT, the provision of nicotine in NVP aerosol reduces cravings and withdrawal symptoms. With practice, NVP use can deliver similar blood nicotine levels to those from cigarette smoking. [23] Weighing up all the available safety data, a 2018 committee report by the US National Academies of Science, Engineering and Medicine concluded that substituting NVPs for combustible tobacco cigarettes reduces a person’s exposure to many toxicants and carcinogens present in combustible tobacco cigarettes and may result in reduced adverse health outcomes. [24] Since that report, the latest Cochrane living review on e-cigarettes for smoking cessation has found that quit rates were higher with NVPs compared to: non-nicotine e-cigarettes; to NRT; and to behavioural support only or no support, with no evidence of harm with up to 2 year follow-up. [25] While NVPs may sit lower on the risk continuum, they are not entirely harmless and require further research.[24, 25]

Large scale population surveys in the US (n=161,054) and UK (n= 170,490) show that NVP use is significantly associated with smoking cessation. [26, 27] The most recent randomised contolled trial (RCT; n = 1124) compared 14 weeks of 21 mg patches alone, 21 mg patches plus NVPs, and 21 mg patches plus nicotine-free e-cigarette. At 6 month follow-up, those in the patches and nicotine e-cigarette group had a significantly higher quit rate (7%) than those in the patches and nicotine-free e-cigarette (4%). The abstinence rate in the patches alone group was 2%, which was not significantly different from the other groups. [28] These quit rates appear to be low, but self-reported point prevalence abstinence rates at 1 month and 3 months were considerably higher and similar to other NRT trials.

Research to date with NVPs for smoking cessation has been mainly conducted in the general population. However, results from small pilot studies suggest the potential benefit of NVPs among people with AOD dependence. [29] We conducted a pilot trial of the current intervention to assess feasibility of implementation, acceptability, safety and potential impact for people being discharged from a smoke-free residential withdrawal unit. [30] The aim of intervening at discharge was to capitalise on the short stay (about 7 days) in the residential unit which is smoke-free, and where most consumers receive NRT to assist them through the smoke-free period. Typically, no support to stay smoke-free is provided after people leave treatment, causing most consumers to immediately return to smoking. This has been a missed opportunity to reinforce stop smoking behaviours. ‘QuitNic’ was a pragmatic two-arm RCT. At discharge from a residential withdrawal, 100 clients received telephone Quitline, free government funded behavioural support service and either 12-weeks supply of NRT or an NVP (plus one week of patches). At 12 weeks, 68% of the NRT group reported using combination NRT while 96% of the NVP group used the device. Acceptability ratings for the products were high in both conditions. At 12 weeks, 14% of the NVP group and 18% of the NRT group reported not smoking at all in the last seven days. Mean cigarettes per day among those who smoke decreased significantly between baseline to 12 weeks in both groups; from 19.91 to 4.72 for the NVP group (p < .001) and from 20.88 to 5.52 in the NRT group (p < .001). Cravings and withdrawal symptoms significantly decreased for both groups. This pilot study showed that smoking cessation support involving options for NRT and Quitline-delivered cognitive-behavioural counselling is attractive to people after they have been discharged from AOD residential withdrawal treatment. Both nicotine vaping products and pharmaceutical NRT were highly acceptable and associated with reductions in cravings for cigarettes, perceptions of withdrawal symptoms and reductions in the number of cigarettes smoked. Some participants reported abstinence from cigarettes; around one in five reported having quit smoking cigarettes at 12 weeks post-discharge. The current trial, Project NEAT, builds on these promising results.

The primary aim is to examine, using a RCT, the effectiveness of interventions consisting of NVPs or NRT plus Quitline counselling on self-reported 6 months continuous abstinence at the 9-month follow-up (6 months following end-of-treatment) for people leaving smoke-free residential withdrawal treatment. Secondary aims are to compare the following outcomes between conditions:

1. Self-reported 30-day point prevalence abstinence at months 3 (end-of-treatment) and 9 (6 months following end-of-treatment)
2. Biochemically verified 6-month continuous abstinence at 9-months follow-up
3. Reduction in cigarettes per day at months 3 and 9
4. Reduction in the frequency and strength of urges to smoke tobacco at months 3 and 9
5. Nicotine withdrawal symptoms in the last 30 days at months 3 and 9

In addition, process measures, including qualitative interviews, will be collected to assess how pharmacotherapies and the Quitline are used by this sample, the barriers and facilitators to making and sustaining a quit attempt, the perceptions of staff at participating sites, as well as Quitline staff, regarding the acceptability and feasibility of the interventions.

## METHODS

### Study Design

The study is a two-arm, single-blinded, parallel-group randomised trial with a 6-month post-intervention follow-up (9-months following baseline).

### Setting

The study will be conducted in six residential and inpatient AOD withdrawal units across four cities in three states of Australia (New South Wales, Queensland, Victoria). All of the sites are smoke-free government-funded or managed health services.

### Participants

Participants may be included in the trial if they meet the following criteria:

- Participating service user aged 18 years or over
- Smoke 10 or more tobacco cigarettes daily on entering withdrawal unit
- Receive NRT while undergoing treatment at participating AOD residential withdrawal services
- Want to quit smoking in the next 30 days
- Have the capacity to consent and able to understand the participant materials and follow the study instructions and procedure (e.g. sufficient English language ability and not too unwell as judged by medical staff)

People will be excluded if they report being pregnant or are breastfeeding, being enrolled in another smoking cessation study, planning to transfer to a long-term residential rehabilitation service following discharge from the withdrawal unit, using a NVP (containing nicotine) in the last 30 days, currently engaging in Quitline’s call-back services, receiving prescribed stop smoking medication (e.g. varenicline or bupropion), or with limited access to a telephone.

### Screening, Recruitment, Baseline Survey

During intake assessment, withdrawal unit staff will notify all service users that the site is participating in a trial about quitting smoking and ask if they would like to discuss the study further with the site research assistant (RA). Prospective participants will be referred to the RA. The RA will complete a formal screening process approximately 1-3 days post intake, explain the project, answer any questions, provide written participant information sheets and obtain written consent. As part of the consent form, participants will be asked to provide their consent for Quitline to call them and to share administrative call data (number, length and timing of calls with Quitline) with the research team. Consent will also be collected for the research team to use participant contact details at the end of the trial to invite the participant to participate in a one-off telephone interview about their study experience. After formal enrolment into the trial, participants will be assigned a unique study ID number in a re-identifiable format. The ID number will be used to enter the secure online survey platform. Participants will first enter their identifying information (follow-up contact details) and then proceed to the baseline questionnaire items administered using an electronic tablet device. The participant identifying details will be stored separately to other data; the ID number will be attached to both.

### Randomisation and Data Management

Independent statisticians from the Statistics Support Unit at the Hunter Medical Research Institute (HMRI) will manage the randomisation service, develop the data management systems (including a quality assurance plan), and conduct statistical analyses. Randomisation will be independently designed and managed using REDCap’s (Research Electronic Data Capture) randomisation module. [31]At the end of the baseline survey, participants will be randomised to an intervention condition via a computer sequenced block randomisation embedded in the electronic tablet. Randomisation in permutated blocks of 4 or 6 will be conducted on a 1:1 allocation basis, stratified by service site. Data will be stored in a REDCap database hosted on HMRI secure servers.

### Randomly assigned groups

The two intervention conditions have been selected based on their potential to assist people who are heavily dependent on smoking cigarretes in converting to a period of abstinence (while in a smoke-free AOD treatment facility) and into sustained cessation (following discharge).

#### Group 1: Current best-practice NRT (comparator) group

will receive 12 weeks supply of dual NRT. NRT has proven effectiveness and few known adverse events. [32] Dual NRT is recommended best practice for people who are heavily dependent smokers and involves coupling sustained-release nicotine patches (e.g., 21mg/24 hours) with fast-acting products, such as nicotine gum or nicotine inhalators, that provide an immediate effect. Research has shown that people who smoke from high-risk groups often cite the cost of NRT as a barrier to use. [33] Thus, the provision of 12 weeks supply free of charge will support treatment adherence. Participants in this group will initially receive four weeks supply of patches plus their choice of either gum or inhalator. They will be asked for a mailing address so that the remainder can be securely couriered to them on two further occasions (3 × 4 weeks supplies).

#### Group 2: Vaporised Nicotine Products NVP (intervention) group

Participants in Group 2 will be provided with an NVP kit including a refillable tank style device (Innokin® Endura T18-II and enough liquid nicotine to last for four weeks (8 × 10ml bottles). Further supplies of nicotine liquid will be delivered via registered post at four-weekly intervals (over the 12 weeks), as per QuitNic pilot study procedures. [30] Participants in Group 2 will be told that they will be sent nicotine liquid refills but no other devices, to minimise on-selling. The Endura T18-II devices meet manufacturing Quality Assurance requirements, are not produced by a company linked to the tobacco industry and were rated high for ease of use and aesthetic appeal by our pilot study participants. As in our pilot study, [30] around the time of discharge, the study RA will provide Group 2 participants with a brief information session on how to use the NVP, along with written materials for safe and effective use to take home. Participants will be advised to use NRT patches (21mg/ 24 hour, 1 week supplied) and the NVP, for the first week following discharge. The liquid nicotine (Nicophar® brand) is manufactured to Good Manufacturing Practice (GMP) standards. [34] It will be provided to participants in 10ml dropper bottles. Each 12mg strength 10mL bottle will contain nicotine (1.2%), Glycerol (84%) and water (14.8%).

#### Behavioural support

Both groups will receive printed information regarding their intervention products. To increase the likelihood of quit success, participants in both conditions will also receive a proactive referral to telephone Quitline support. Research has repeatedly shown that a combination of pharmacotherapy and behavioural support results in higher cessation rates than either approach alone. [35] All participants will be referred to the Victorian Quitline as a central line for all study participants. There is strong evidence of the effectiveness of proactive telephone calls for smoking cessation (e.g. Cochrane review, n = 24,000, reporting relative risk of 1.37 at longest follow-up). [36] However, research also suggests that people who smoke from high-risk groups are less likely to call quitlines. [20] Proactive approaches (arranging for Quitline to call the smoker) are important to increase engagement with the service. For this study, Quitline counsellors will be trained to provide cessation support to people with substance use disorders, as was done in the QuitNic pilot study. [30] In the current study, the first call will occur while in residential care with post-discharge calls focusing on relapse prevention. Zhu’s relapse prevention call schedule [37] will be followed with calls on days 1,3,7,14 and 28 post-discharge. The total number and timing of calls will be tailored to service user need and smoking status (i.e., more frequent calls around relapse crises/quit attempts) with a maximum of 15 calls over 12 weeks. Participants will be sent a text message prior to being called to encourage answering calls from a private number (Quitline). Counsellors will monitor and promote the correct use of NRT and NVPs and address barriers to their use.

### Blinding

All outcome assessments will be conducted by the independent research company using computer-assisted telephone interviewing (CATI). The interviewers who conduct outcome assessments will use the unique study ID (i.e. randomisation number) and will not know the person’s treatment allocation. The outcome assessors will not collect any adverse events data, which will be done by a separate group of interviewers. The investigators will be blinded to participants’ treatment allocation until all data have been collected.

### Minimising contamination

There is a possibility that participants within a service randomised to one condition (e.g., NRT) will be exposed to the other condition (e.g., NVPs). The study design reduces the risk of contamination: although participants will be trained in the use of NVPs while in the service, they will not receive the NRT or NVP packs until they are discharged from the facility. Following discharge, our pilot study indicates that participants are unlikely to see each other. Our pilot trial also found no evidence of contamination occurring, with only two participants in the NRT group expressing interest in NVPs but compliant with their allocation. Use of other types of cessation aids, including NVPs, will be assessed in both groups during each follow-up.

### Follow-up and minimising attrition

The study has a 12-week intervention phase, following discharge from the smoke-free facility, to receive and use the supply of NVP or NRT, and Quitline support. Follow-up assessments will be conducted at the end of the intervention phase (3-months) and six months following the end of the intervention (i.e., 9-months following discharge from the AOD service), as per international recommendations for assessing long-term smoking cessation. [38, 39] Follow-up surveys will be administered using CATI software contracted to a research survey company. We will implement retention strategies developed in response to our pilot study [30] to reduce loss to follow-up and according to evidence-based best practice. [40] The CATI company will try to contact each participant up to 10 times at different times of the day and during the week. Participants with mobile phone numbers will be sent reminder text messages each month, asking them to notify the research team if they change their contact details. Participant contact logs will contain the contact details of significant others as well. In addition, loss to follow-up will be minimised by using a $50 reimbursement for the completion of each follow-up survey. All participants reporting abstinence at follow-up will be asked to attend a carbon monoxide test with the site-specific RA and will be given a further $50 reimbursement for travel.

### Outcome Measures

#### Primary outcome measure

The primary outcome will be self-reported 6-month continuous abstinence at 9-months post-discharge, not having smoked more than five cigarettes for the entire 6-month period preceding the follow-up assessment. The primary outcome will be assessed using a composite measure including: i) current smoking status as measured by the item “Do you currently smoke any tobacco products?” with response options Yes, daily, Yes, at least once a week, Yes, less often than once a week, No, not at all; ii) duration since quitting smoking “How long ago did you finally stop smoking everyday?” with response options Less than a week, 1 week to 1 month, 1-3 months, 4-6 months, More than 6-months; iii) occasions of smoking since quitting as measures by the item “Since you quit smoking, have you smoked every day for a week or more (even just puffs)?” with response options yes, no. These items together will inform whether individuals meet self-reported 6-month continuous abstinence from tobacco smoking.

#### Secondary outcome measures

have been pilot tested in the QuitNic trial and constructed to be brief but contain all necessary measures. Collected at months 3 and 9 will include self-reported *30-day point prevalence smoking abstinence*, based on, “Have you smoked at least part of a cigarette in the last 30 days?”; [39] *point prevalence abstinence* from all nicotine (including NRT and NVPs): defined as having not used any products containing nicotine in the previous 30 days at assessment; *cigarettes smoked per day*; *frequency of cravings and strength of urges*: assessed by items based on Taggar et al. [41] Currently, how often do you get strong cravings to smoke tobacco products?” with the response options of 1) Hourly or more often; 2) Several times per day; 3) At least once a day, 4) Less than daily, 5) Never and “How strong have these urges been? 1) No urge; 2) Slight; 3) Moderate; 4) Strong; 5) Very strong; 6) Extremely strong; *withdrawal*: as assessed by the Minnesota Nicotine Withdrawal Scale, [42] An eight-item scale rating severity of withdrawal symptoms on an ordinal scale ranging from 0 (not present) to 3 (severe); *time to relapse*: will be measured using Ursprung et al.’s Latency to Needing a Cigarette Scale; [43] *number of subsequent quit attempts* will be assessed among those who relapsed.

##### Biochemical verification of self-report

Self-reported 30-day point prevalence abstinence at 6-months post-treatment will be verified using a measure of carbon monoxide (CO) in expired air. [39, 44] Twenty-four hours is the recommended biochemically verifiable window for CO in expired air. To assess 24-hour smoking cessation, participants will be asked how long ago they last smoked a cigarette, cigar or pipe? (respond by indicating, last 4 hours, 4-8 hours ago, 9-24 hours ago, > 24 hours ago). The Jarvis protocol will be used to record expired air CO. A cut-off point of 8 ppm will be used to indicate that recent smoking has occurred. [44] Measuring CO in expired air has demonstrated high sensitivity and specificity of around 90% for both. [44] We will conduct secondary analyses reporting on the proportion of self-reported abstinence.

#### Other measures

Several potential baseline covariates, mediating factors, process measures and other outcome variables will also be collected at baseline and each follow-up. Adverse events will be recorded and coded for seriousness, severity, causality and expectedness. Table 1 lists the data collection schedule for all variables (primary, secondary, covariates, mediating factors and exploratory outcomes).

**Table 1.**
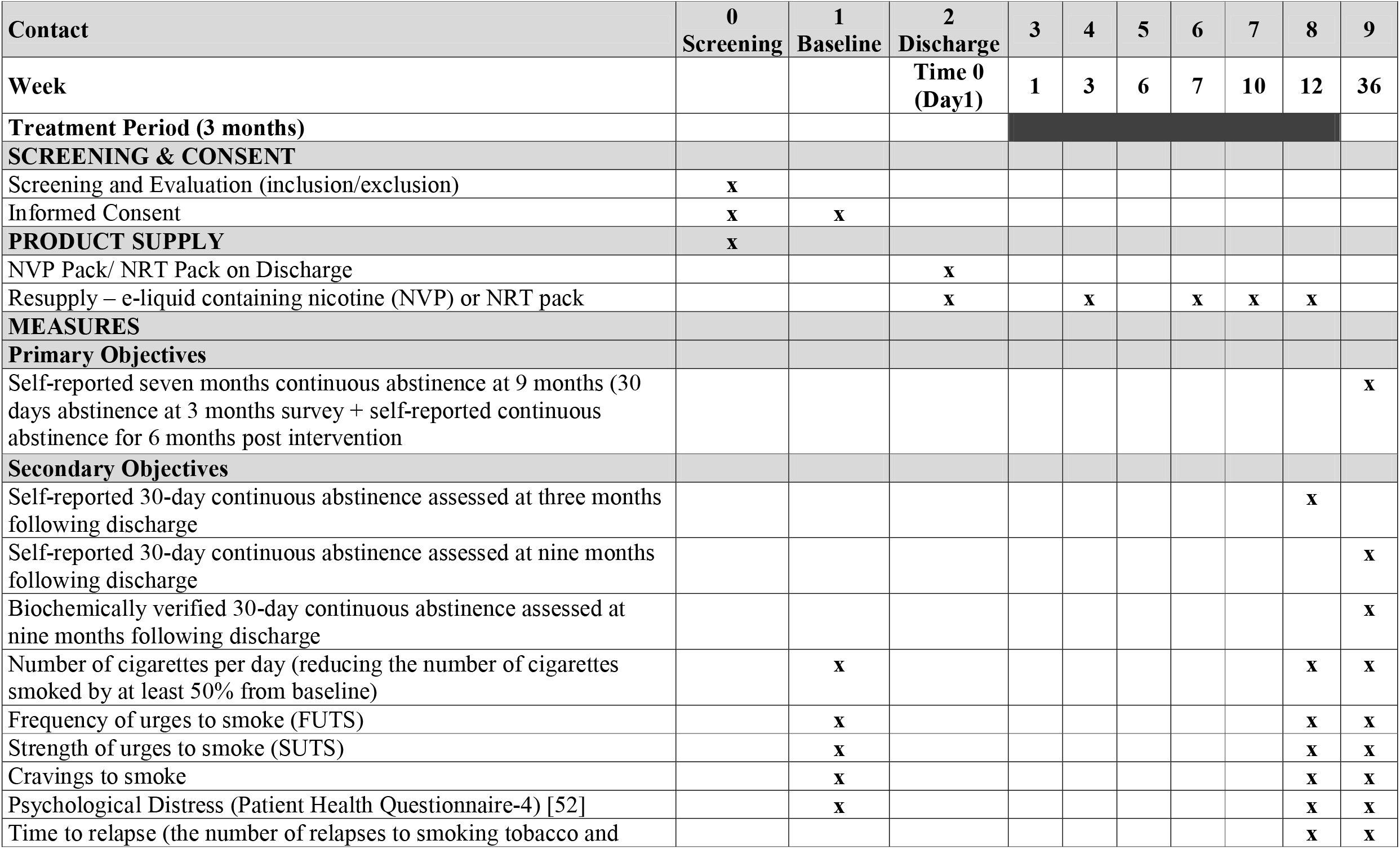
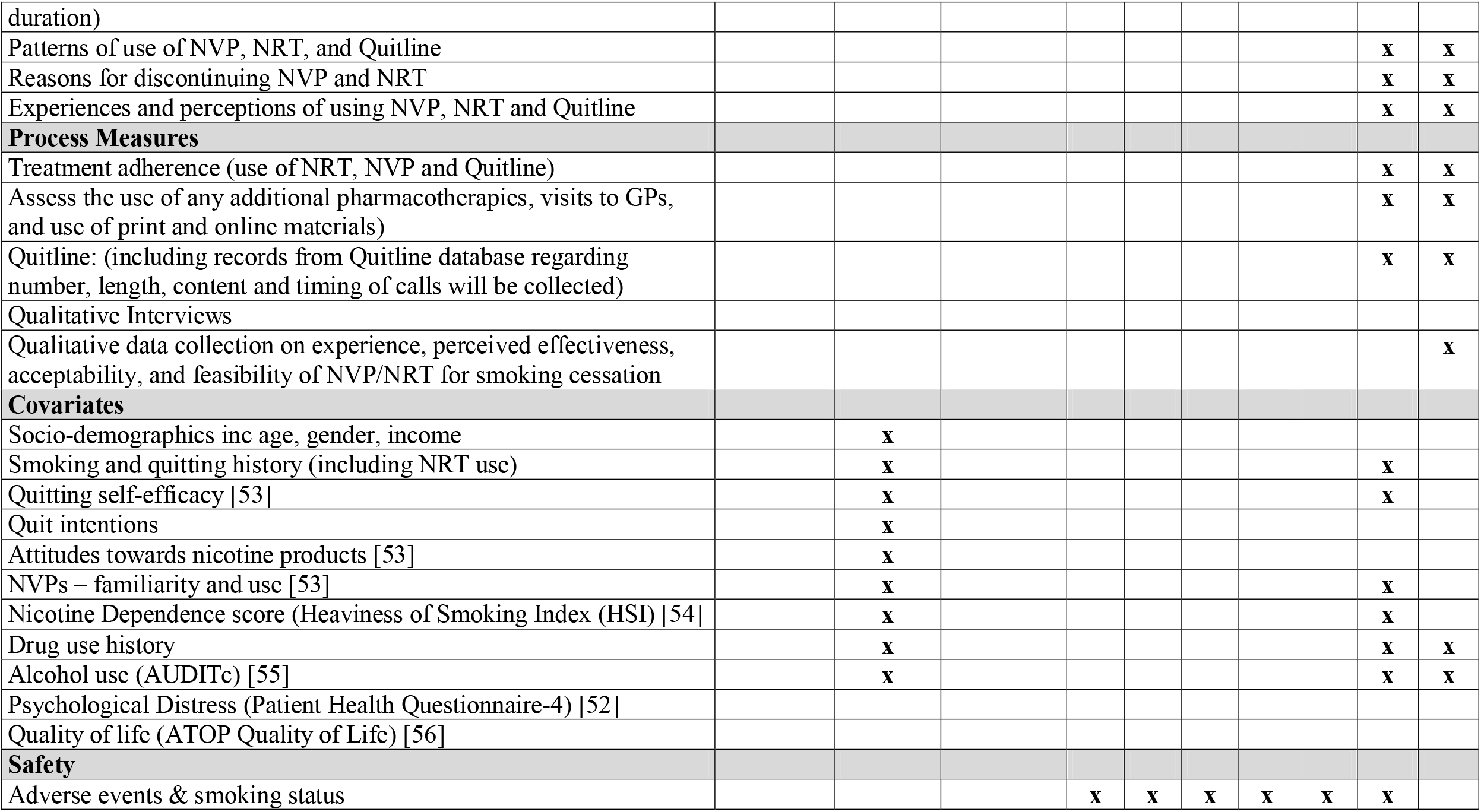
Schedule of activities.

### Statistical Methods

The analysis plan will be pre-specified. Descriptive statistics will be used to present the baseline characteristics for intervention and control groups. All statistical tests will be two-tailed and conducted at the 5% significance level.

#### Aim 1

The primary outcome of interest will be the proportion of people reporting abstinence at nine months post-randomisation measured using 6-months continuous abstinence measure. Differences between groups will be assessed using logistic regression. The model will include a term for treatment group while adjusting for potential confounders, including service site. Odds ratios will be calculated, together with 95% confidence intervals and likelihood ratio test p-values. As recommended by the Russell Standard, [39] the primary analysis will be intention-to-treat (ITT); all consenting participants in the cohort will be included in the analysis. Those lost to follow-up for any reason will be regarded as smoking. Sensitivity analysis will be performed using multiple imputation modelling and appropriate techniques if the data are considered to be missing not at random (pattern mixture models, for example).

#### Secondary aims

For the secondary outcomes, a logistic regression model for abstinence outcomes will be used (continuous abstinence at 3-months, self-reported point prevalence abstinence at 3- and 9-months) unadjusted for other covariates. The number of cigarettes smoked per day will be compared at each time point using a negative binomial regression model. Craving and withdrawal scores will be compared using ordinal logistic regression models. Change from baseline in each of the repeated measures and cigarettes smoked per day (in non-abstainers) will be analysed using linear mixed models with a compound symmetry covariance structure.

##### Sample size

Studies using continuous abstinence measures with mental illness and AOD samples have found smoking cessation rates close to zero at longer-term follow-up using NRT. [45] Using the same outcome measures, Bullen and Walker [46] found no significant difference in continuous abstinence between NVP and NRT groups (7.3% vs 5.8% respectively at six months follow-up) in a general population sample in NZ. The abstinence rates in the QuitNic pilot trial in a sample of AOD treatment service users were similar for both NVP (14%) and NRT (18%), but higher than the results found by Bullen and Walker. However, the measure used in the QuitNic pilot was the much less conservative 7-day point prevalence abstinence measure, assessed only at end-of-treatment (12 weeks). Given the current trial is using a long-term follow-up and continuous abstinence measure, we will conservatively assume a continuous abstinence rate of 3% at six months follow up in the NRT group. A sample of 278 people who smoke in each treatment group is needed to detect a difference of 6% between groups (i.e., 3% in NRT group and 9% in NVP group continuous abstinence at 9-month follow-up) with 80% power and a 5% significance level.

### Clinical trial governance

University of Newcastle was the trial sponsor to March 2021 and was transferred to Flinders University under a clinical trial Collaborative or Cooperative Research Group agreement when the Principal Investigator (Bonevski) relocated. The trial is registered in the Australian and New Zealand Clinical Trials Registry (ACTRN12619001787178). We will establish a Protocol Steering Committee (PSC) consisting of Investigators and staff, which will meet regularly via teleconference in early phases of the trial to monitor progress and resolve any issues that arise. This committee will distribute progress reports and seek input from the wider team at key progress points and as needed. The research team will send regular newsletters to trial sites and stakeholders.

#### Regulatory Approvals and Compliance

The sponsor (Flinders) will supply the unapproved therapeutic goods (nicotine vaporiser and refill solution) under a Therapeutic Goods Administration (TGA) Clinical Trial Notification, and comply with Good Clinical Practice (GCP) and the NSW Health Poisons and Therapeutic Goods Regulation 2008 under the Poisons and Therapeutic Goods Act 1966, which authorises the supply of restricted drugs for clinical trials.

#### Participant safety and adverse events

We will follow the NHMRC and TGA Guidelines for monitoring and reporting in clinical trials involving therapeutic goods. [47, 48] A number of team investigators are providing clinical oversight. We will establish an independent Data Safety Monitoring Board (DSMB) comprised of an experienced statistician and medical experts with specialist training in addiction medicine. The DSMB will provide oversight of safety parameters, methods and timing of assessment; recording and reporting serious adverse events and inter-current illnesses; the type and duration of participant follow-up required after adverse events. The DSMB will set clear stopping rules and be guided by statistical monitoring guidelines, consistent with GCP. The Protocol Steering Committee will provide the DSMB with a report for each meeting detailing participant recruitment, number of withdrawals, side effects and any serious or unexpected adverse events both related and unrelated to the provided investigational products. The DSMB will meet according to a schedule established prior to study initiation and at other time-points according to perceived need. We will report all serious adverse events (related and unrelated) to the lead human research ethics committees (Hunter New England HREC and The University of Newcastle) using their SAE reporting forms. These reports will be sent to the research governance offices (RGOs) provide site specific authorisation for the participating trial sites. Adverse events will be included in annual reports to the HRECs. We will record adverse events in GCP compliant software. All suspected unexpected serious adverse drug reactions (SUSARs) as well as unanticipated serious adverse device effects (USADE) will be reported to the HRECs and RGOs, and the investigators as soon as possible and to the TGA Drug Safety and Evaluation Branch, according to the TGA reporting guidelines.

### Dissemination policy

Results will be disseminated to key stakeholders, and more broadly, in several ways. These include open-access or pre-print peer-reviewed journal articles; presentation at scientific meetings; feedback to trial participants and participating sites; and press releases. De-identified individual participant data and data dictionaries will be available beginning 3-months and ending 5-years after the primary outcomes manuscript is published. Proposals (using a proforma created by the study team and provided upon request) are to be submitted to the coordinating principle investigator (Bonevski) for access to the data. Analysis files will be provided with the published primary outcomes manuscript. The criteria for authorship will be taken from the International Committee of Medical Journal Editors. [38]

### Ethics

The trial will be conducted in compliance with the protocol, the principles of Good Clinical Practice, [49] the National Health and Medical Research Council’s (NHMRC) National Statement on Ethical Conduct in Human Research (2007) [48]. Ethical approval for this study has been obtained from Hunter New England Area Health Service (REGIS: 2019/ETH10554) and the University of Newcastle Human Research Ethics Committee (H-2019-0358) and site-specific approvals from each participating site. A TGA clinical trial notification has been filed as the Nicotine Vaping Product is not a registered product listed on the Australian Register of Therapeutic Goods (ARTG).

## DISCUSSION

Addressing tobacco use in populations with the highest smoking prevalence is an international priority. High smoking rates amongst people who use other substances undermine their treatment success and general health and contribute to high rates of premature mortality. Time in a smokefree AOD residential treatment facility is an ideal opportunity to commence smoking cessation support to quit smoking. Clients may be living in the smoke-free facility for days or weeks, however the majority of clients relapse back to smoking upon discharge from treatment. This study will use the window of opportunity during discharge to offer clients two forms of nicotine-based smoking cessation support, plus telephone counselling. This is the first study we know of that will be testing the effectiveness of NVPs in combination with telephone counselling support during this transition phase from treatment to community. From a public health perspective, this approach also has the potential to have tremendous reach into a priority population for smoking cessation with over 220,000 treament episodes to an estimated 137,000 clients each year in Australia. [3] There is also potential to scale up this intervention in private providers of drug and alcohol treatment services. Most of these clients smoke tobacco cigarettes.

Despite the burden that tobacco places on people with AOD dependence, there are no effective interventions for achieving long term smoking abstinence, and quit rates are close to zero. This trial is testing an innovation in replacement nicotine delivery at a time in a person’s treatment journey that it is likely to be beneficial. NVPs hold significant potential as smoking cessation treatment, potentially reducing relapse to smoking and need to be tested amongst this group. There is currently no other adequately powered RCT published of NVPs for smoking cessation support amongst people seeking treatment of AOD dependence. This trial will provide invaluable data for shaping policy in Australia and world-wide regarding the use of NVPs and the optimal setting for implementation. In addition, the trial will also provide new knowledge on the uptake and effectiveness of combination NRT for smoking cessation among AOD service users.

There continues to be some debate regarding the safety of NVPs with data suggesting they are harmful products coming from predominantly in vitro studies or clinical case studies reporting small numbers of burns, suspected acute lung injury or acute nicotine toxicity. [50] There is no evidence linking e-cigarettes to the highest burden diseases caused by or linked to tobacco smoking including cardiovascular disease, cancer, respiratory conditions, mental health, development in children and adolescents, reproduction, sleep, wound healing, neurological conditions other than seizures, and endocrine, olfactory, optical, allergic and haematological conditions. [50] Furthermore, all randomised controlled trials of NVPs have failed to identify safety issues. [25] Although not a safety trial, this trial will add to the evidence base of the safety of NVPs with reporting of adverse events in an otherwise high morbidity group.

Finally, detailed data will be collected from all participants on their use of telephone Quitline throughout the study. Quitline quit specialists (counsellors) will be given bespoke training to improve engagement with AOD service users. Booster training will be provided by the trial coordinating centre with monthly supervision provided by a senior Quitline staff member. This study will provide data on the acceptability of this tailored form of telephone counselling.

Strengths of this study are the large sample size, multiple settings and rigorous study design. The trial was developed with AOD clinician and consumer involvement and following a high-quality pilot trial. [30] The telephone-based interviews and assessments will improve the recruitment and retention rate, as it will allow greater flexibility for participants to conduct interviews. A suite of retention strategies will be used to minimise attrition rates, an issue identified in the pilot study. Another strength is that self-reported smoking cessation for 30-day point prevalence at the 9-month follow-up will be verified using CO breath analysis. It is impossible to biochemically verify 6-month continuous abstinence (the primary outcome) in this sample of other substance treatment seekers. Use of the CO biochemical verification will provide a proxy measure of the accuracy of self-report. There are limitations in using CO breath analysis as cannabis use, usually high in this sample, is detected in the breath analysis. However, since some participants may continue using their nicotine replacement products at our follow-up points, it was also not possible to use cotinine measures which detect nicotine. We have included questions in our follow-up survey regarding cannabis smoking to assess the likelihood of it affecting CO breath analysis results.

This research has significant potential to influence policy and practice. Australian policy supports NVP use as a smoking cessation aid if approved by TGA as a therapeutic good. The Royal Australia College of General Practitioners have recommended use of NVPs for smoking cessation as a second line treatment if other evidence-based treatments are ineffective. [51] The method of prescribing and dispensing liquid nicotine and the NVPs within health services, under clinical oversight, is a model that aligns with Australia’s regulatory framework. Hence, clinical trial evidence on their efficacy and safety, such as will be collected in this study, is critical. The trial is conducted collaboratively with key stakeholder organisations in Australia with an interest in this work (e.g. NSW Health, Queensland Health, VicHealth, Victorian Quitline) and will work closely with the Project Advisory Committee to translate the results into policy recommendations.

### Clinical trial registration

The trial has been prospectively registered with Australian and New Zealand Clinical Trials Registry (ACTRN12619001787178). The trial started recruitment on the 28^th^ September 2020, with a nine-month delay caused by the Covid-19 global pandemic outbreak. Recruitment is expected to take 24 months, and trial findings are scheduled to be reported in late 2023

## Data Availability

Data is currently being collected and may be available upon reasonable request to the authors

## Acknowledgements

This study is funded by an Australian National Health and Medical Research Council (NHMRC) Project Grant (GNT1160245). The authors thank Edwina Williams for her research assistance.

